# Real-World Effectiveness and Safety of GLP-1 Receptor Agonists in Patients with Weight Recurrence After Bariatric Surgery

**DOI:** 10.1101/2025.11.21.25340747

**Authors:** Tarik Yuce, Rotana M Radwan, Yao An Lee, Emma Holler, Hao Dai, Xing He, Yu Huang, Yi Guo, Jiang Bian, Jingchuan Guo

**Affiliations:** Surgical Outcomes and Quality Improvement Center (SOQIC), Department of Surgery, Indiana University School of Medicine, Indianapolis, IN, USA; Department of Pharmaceutical Outcomes and Policy, College of Pharmacy, University of Florida, Gainesville, FL, USA; Department of Pharmacy Practice, Purdue University College of Pharmacy; Department of Biostatistics & Health Data Science, Indiana University, Indianapolis, IN, USA; Center for Biomedical Informatics, Regenstrief Institute, Indianapolis, IN, USA; Department of Health Outcomes and Biomedical Informatics, College of Medicine, University of Florida, Gainesville, FL, USA

## Abstract

**Importance:** Weight recurrence occurs in nearly 20% of patients following metabolic and bariatric surgery (MBS) and can diminish long-term metabolic benefits. While glucagon-like peptide-1 receptor agonists and gastric inhibitory polypeptide receptor agonists (GLP-1 RA) have emerged as potential adjuvant therapies, evidence supporting their use is limited primarily to liraglutide with minimal real-world data on newer agents or population-level effectiveness.

**Objective:** To evaluate the effectiveness and safety of GLP-1 RA therapy compared to non-use among patients experiencing weight recurrence following MBS.

**Design, Setting, and Participants:** This retrospective new-user cohort study utilized from the OneFlorida+ Data Trust (2015-2024) to identify adults who underwent MBS and subsequently experienced ≥10% weight recurrence from nadir weight. Propensity score matching based on BMI and clinical encounter timing, combined with inverse probability of treatment weighting, was used to minimize confounding. Of 1,098 patients who underwent MBS and experienced weight recurrence, the final weighted analytic cohort included 68 GLP-1 RA users and 131 non-users at 6 months, and 72 users and 139 non-users at 12 months.

**Exposure:** Initiation of any GLP-1 RA (dulaglutide, exenatide, liraglutide, lixisenatide, semaglutide, or tirzepatide) after MBS among patients with weight recurrence.

**Main Outcomes and Measures:** The primary effectiveness outcome was percentage of recurrent weight lost at 6 and 12 months. Safety outcomes included composite gastrointestinal and surgical adverse events.

**Results:** GLP-1 RA use was associated with significantly greater mean weight recurrence loss compared to non-use at 6 months (9.6% vs 4.3%, p = 0.005) and 12 months (14.8% vs 8.1%, p = 0.017). Among individual agents, tirzepatide was associated with the largest reductions at both 6 and 12 months (25.1% vs 5.4% and 34.4% vs 8.8%, respectively; both p < 0.001). Liraglutide demonstrated a statistically significant difference at 12 months (18.3% vs 8.4%, p = 0.004), while semaglutide and dulaglutide were not associated with statistically significant differences at either time point. No significant differences in adverse event rates were observed between groups, with composite safety outcomes occurring in 47.1% of users versus 56.5% of non-users at 6 months (HR = 1.24, 95% CI: 0.92-1.68, p > 0.05).

**Conclusions and Relevance:** Use of GLP-1 RA for the treatment of weight recurrence following MBS represents an effective option with sustained benefits through 12 months without increased risk of adverse events. These real-world findings support adjuvant GLP-1 RA use for managing weight recurrence following MBS across diverse patient populations.

**Key Points:** *Question:* Does the use of glucagon-like peptide-1 and gastric inhibitory polypeptide receptor agonists (GLP-1 RA) for the treatment of weight recurrence after bariatric surgery lead to significant weight loss in real-world populations?

*Findings:* In this cohort study of 1,098 individuals who underwent bariatric surgery and experienced weight recurrence, use of GLP-1 RA was associated with significant loss of regained weight.

*Meaning:* Use of GLP-1 RA for weight recurrence after bariatric surgery represents a viable treatment option.

## Introduction

Obesity continues to represent a growing health care burden globally.^1,2^ Metabolic and Bariatric surgery (MBS), though underutilized, represents one of the most powerful treatments for obesity to date.^3–6^ While patients experience substantial weight loss following MBS, nearly 20% experience weight recurrence defined as _≥_10% of nadir weight.^7,8^ The FDA approval of novel anti-obesity medications, including glucagon-like peptide-1 and gastric inhibitory polypeptide receptor agonists (collectively GLP-1 RA), for weight loss has led to increased interest in understanding if and how these medications can be used in conjunction with MBS (i.e., neoadjuvant, perioperative, adjuvant).^9–12^

While the adjuvant use of GLP-1 RA to treat MBS weight recurrence has become more common, the evidence supporting this approach is limited.^13^ While two recent clinical trials have supported the use of adjuvant liraglutide to treat weight recurrence or poor weight loss following MBS, comparative data for newer GLP-1 RA (e.g., semaglutide, tirzepatide) are limited to single-center observational studies.^14,15^ As a result, there is minimal high-quality evidence to guide the selection of GLP-1 RA for treatment of weight recurrence outside of liraglutide. Furthermore, real-world evidence supporting the use of these treatments across diverse populations and settings remains limited given the small cohort sizes and single-institutional nature of most current studies.^16–19^ These evidence gaps highlight the need for population-level studies using causal inference methods to inform clinical decision-making regarding selection and use of adjuvant GLP-1RA for MBS weight recurrence.

Given the growing prevalence of GLP-1 RA use following MBS, there is a critical need for rigorous real-world studies evaluating the effectiveness and safety of this approach. This study addresses these gaps by combining population-level electronic health record (EHR) data with target trial emulation methodology to provide guidance for the adjuvant use of GLP-1 RA for MBS weight recurrence. The objectives of this study were to: (1) compare the effectiveness of GLP-1 RA initiation following MBS to non-use for the treatment of weight recurrence, (2) evaluate whether treatment effects vary by individual GLP-1 RA or patient characteristics, and (3) determine the comparative safety of GLP-1 RA use in this population.

## Study Data and Methods

### Study Design and Data Source

This retrospective new-user cohort study evaluated the effectiveness and safety of GLP-1RAs use compared with non-use among adults (_≥_18 years old) who had recurrence of at least 10% of their nadir (lowest) weight following bariatric surgery. Data were obtained from the OneFlorida+ Data Trust, a centralized patient data repository that integrates longitudinal electronic health records (EHRs) with additional sources such as Medicaid and Medicare claims, death records, and tumor registry data.^20^ As of 2025, the repository contains data on approximately 26 million individuals, with records dating back to 2012. The database includes a wide range of patient characteristics, including demographics, diagnoses, prescribed medications, procedures, vital signs, and laboratory results. The OneFlorida+ Clinical Research Consortium maintains the OneFlorida+ Data Trust, one of eight clinical research networks within PCORnet, the National Patient-Centered Clinical Research Network. This study adhered to the Strengthening the Reporting of Observational Studies in Epidemiology (STROBE) guidelines and was approved by the University of Florida Institutional Review Board.

### Population

The study population included adults (_≥_18 years) who underwent any bariatric surgery between January 1, 2015, and January 31, 2024. The cohort entry date was the first recorded bariatric surgery during the study period. Patients were included if they had at least 12 months of baseline data and a minimum of 6 months of follow-up, with no documented GLP-1 RA use prior to the cohort entry date. Exclusion criteria were pregnancy, active malignant neoplasm, revisional bariatric surgery, and missing baseline body mass index (BMI). International Classification of Diseases, Ninth and Tenth Revision (ICD-9 and ICD-10) and Current Procedural Terminology (CPT) codes used to identify bariatric surgery are presented in **Supplementary eTable 1**.

### Exposure and Follow-up

The exposure of interest was initiation of a GLP-1 RA (i.e., dulaglutide, exenatide, liraglutide, lixisenatide, semaglutide, and tirzepatide) after bariatric surgery. Semaglutide, liraglutide, and tirzepatide are approved for obesity management, whereas the remaining GLP-1 RAs are indicated for type 2 diabetes. Therefore, all GLP-1 RAs were evaluated collectively, with planned subgroup analyses by indication and individual agent.

Patients who initiated a GLP-1 RA were compared with a contemporaneous group of non-users who met the same inclusion and exclusion criteria but did not receive any GLP-1 RA during the study period. For GLP-1 RA users, the index date was defined as the date of first GLP-1 RA prescription following bariatric surgery. For non-users, the index date was assigned by matching each GLP-1 RA user to up to four non-users based on BMI and timing of any inpatient or outpatient encounter within ±4 weeks of the initiator’s index date. This matching aligned both groups on BMI and ensured comparable follow-up time relative to a similar point post-surgery. Patients were followed from the index date until the death, revisional bariatric surgery, loss to follow-up, or end of the study period (January 31, 2024).

### Effectiveness Outcomes

The effectiveness outcome was the percentage of weight recurrence lost at 6 and 12 months after the index date. Weight recurrence was defined as any weight increase occurring after a patient’s lowest recorded post-bariatric surgery weight (nadir) but before the index date (GLP-1 RA initiation for users or the matched assigned index date for non-users). Eligible patients were restricted to those who had recurrence of at least 10% of their nadir weight, representing a clinically meaningful threshold for post-surgical weight recurrence.^8,21^ The percentage of recurrence weight lost was calculated as:

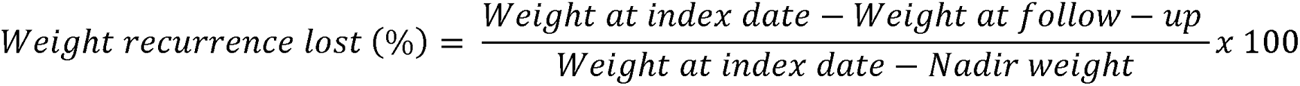

This measure reflects the proportion of recurrent weight that was subsequently lost following the index date.

### Safety Outcomes

The safety outcome was defined as the incidence of adverse events (AEs) following the index date. AEs included abdominal pain, nausea with or without vomiting, gastroesophageal reflux disease (GERD), gastroparesis, dehydration, bowel obstruction, internal hernia, incisional hernia, fistula of the stomach or duodenum, post-bariatric surgery symptoms, and general complications of bariatric surgery. Additional outcomes, such as gastrojejunal ulcer, gallstone disease (cholelithiasis, cholecystitis, cholangitis, and choledocholithiasis), and gastrointestinal hemorrhage, were also evaluated. Safety outcomes were assessed individually and as composite endpoints at 6 and 12 months of follow-up after the index date. ICD-9 and ICD-10 codes for AEs are provided in the **supplementary eTable 2.**

### Covariates

Baseline covariates included demographic characteristics, clinical factors, and comorbidities. Demographic variables were age (18–40, 40–65, and ≥65 years), sex, race/ethnicity (Asian, Black, Hispanic, non-Hispanic White, and Unknown), and insurance type. Clinical variables included baseline BMI (categorized as <25, 25–30, 30–35, 35–40, and ≥40 kg/m²), weight and BMI measurements closest to the index date (including nadir weight and nadir BMI), and time from bariatric surgery to the index date. Smoking status was classified as current, former, or never. Comorbidities included hypertension, dyslipidemia, coronary artery disease, myocardial infarction, stroke, type 2 diabetes, prediabetes, chronic obstructive pulmonary disease, asthma, gastroesophageal reflux disease, gastroparesis, depression, rheumatoid arthritis/osteoarthritis, and sleep apnea.

### Statistical Analysis

We employed a two-stage approach to minimize confounding and improve baseline comparability between GLP-1 RA users and non-users. First, we applied propensity score matching to create a matched cohort. Non-users were matched to users at a ratio of up to 4:1 based on BMI closest to the index date and the timing of a clinical encounter within ±4 weeks of the initiator’s index date. This matching strategy aligned groups on BMI and ensured comparable follow-up opportunities relative to a similar timepoint post-surgery. Second, within the matched cohort, we applied inverse probability of treatment weighting (IPTW) using a propensity score model to further adjust for residual confounding. The propensity score model included sociodemographic characteristics, baseline comorbidities, and concomitant medications to further adjust for residual confounding. Covariate balance was assessed using standardized mean differences, with values less than 0.1 indicating adequate balance after weighting.

For the effectiveness analysis, we compared the percentage of weight recurrence that was subsequently lost at 6 and 12 months between users and non-users. Percentage of weight recurrence (which could include negative values) was right-skewed, thus we applied a signed log1p transformation. Group differences were tested using IPTW-weighted Welch two-sample t-tests on the transformed scale. For interpretability, we report IPTW-weighted arithmetic means on the original percent scale, obtained by back-transforming each individual’s value and then averaging with IPTW weights; p-values reflect comparisons on the transformed (log) scale.^22^ Subgroup analyses were performed according to GLP-1RA type as well as baseline type 2 diabetes status.

For safety outcomes, we calculated cumulative incidence proportions at 6 and 12 months for the pre-specified adverse events of interest, both individually and as a composite. Risk was evaluated using Cox proportional hazards models applied to the IPTW cohort, generating hazard ratios (HRs) and 95% confidence intervals (CIs) comparing GLP-1 RA users to non-users. The analysis followed an intention-to-treat approach, where patients were analyzed according to their initial treatment group regardless of treatment discontinuation or switching. All analyses were conducted using Python version 3.10, and statistical significance was defined as a two-sided p-value less than 0.05.

## Results

### Cohort Selection

Between 2015 and 2024, a total of 35,250 patients with a record of bariatric surgery were identified in the OneFlorida+ Data Trust. Among these, 2,629 (7.5%) were GLP-1RA users and 32,611 (92.5%) were non-users. After restricting to adults (≥18 years) with at least one year of baseline data and six months of follow-up, and excluding those without baseline BMI, pregnancy, revisional bariatric surgery, or active malignancy, 284 GLP-1RA users and 9,091 non-users remained. Following time-conditional propensity score matching, the final analytic cohort included 220 GLP-1RA users and 878 non-users.

Weight outcomes were evaluated at 6 and 12 months following the index date. Among these, 71 users and 134 non-users had experienced weight recurrence (i.e., ≥10% of their nadir weight prior to the index date) at 6 months, corresponding to 68 and 131 patients, respectively, after weighting. At 12 months, 74 users and 143 non-users had experienced weight recurrence, corresponding to 72 and 139 patients, respectively, after weighting. The detailed cohort selection process is shown in **Figure 1**.

**Figure 1.**
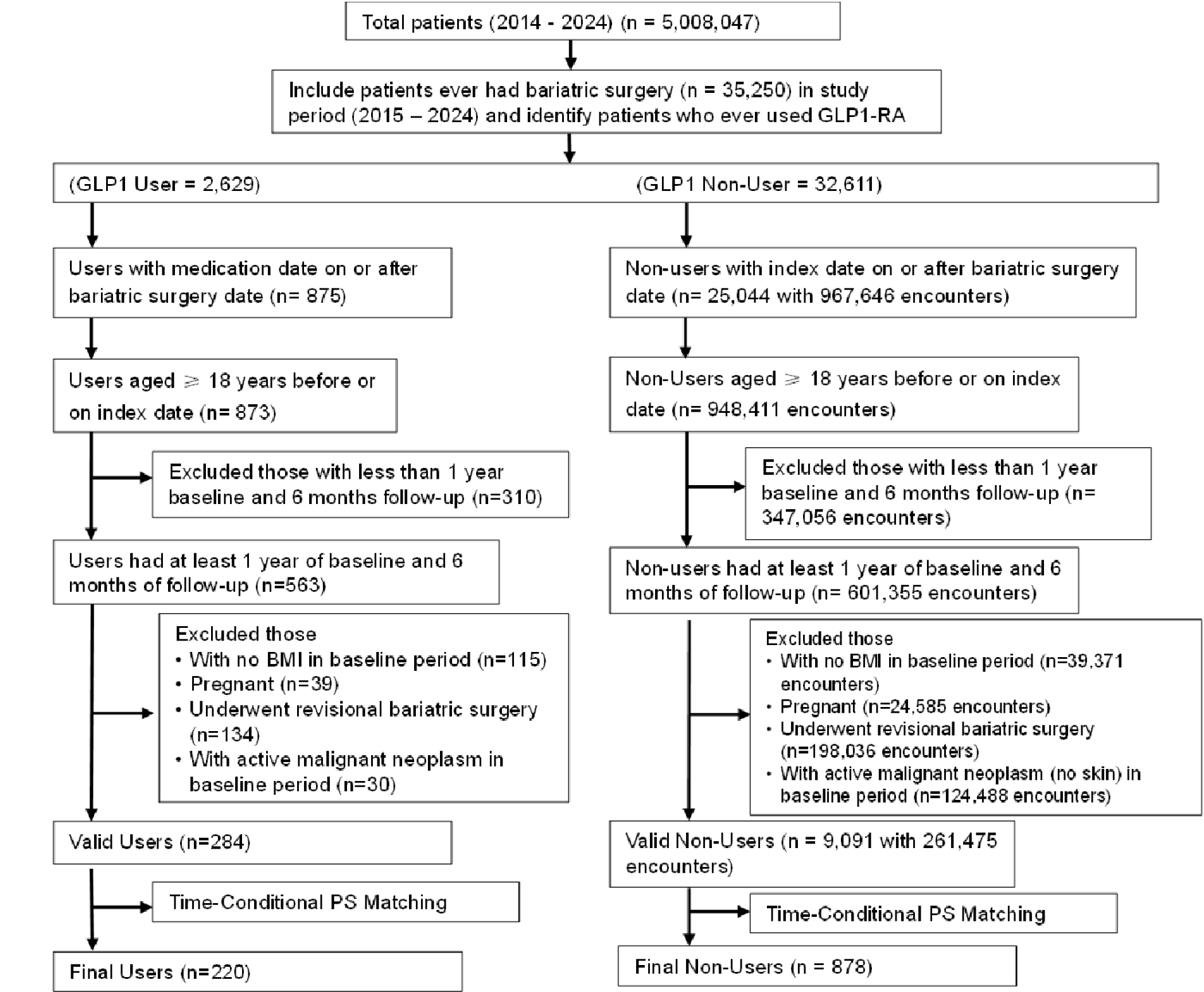
Flowchart of patient selection

### Baseline Characteristics

Before applying IPTW, GLP-1 RA users differed from non-users across several baseline characteristics. Users were younger, more likely to be female, and had higher body weight and BMI at the index date. In the 6-month cohort, mean body weight at index was 248.3 lb (SD = 47.7) among users compared to 237.3 lb (SD = 48.3) among non-users. Mean BMI was 40.7 (SD = 7.9) in users versus 38.3 (SD = 7.7) in non-users. Nadir body weight was 203.6 lb (SD = 43.0) in users and 191.0 lb (SD = 41.3) in non-users, with corresponding nadir BMI of 32.6 (SD = 7.2) and 30.7 (SD = 6.8), respectively. The average time from bariatric surgery to the index date was 49.2 months for users compared to 53.6 months for non-users. Similar baseline imbalances were observed in the 12-month cohort. Similar patterns were observed in the 12-month cohort.

After IPTW, covariates were well balanced between groups in both cohorts (all SMDs <0.1). In the weighted 6-month cohort, mean BMI at index was 40.3 (SD = 7.7) among users and 39.7 (SD = 8.5) among non-users, with comparable nadir BMI (32.1 vs. 31.8) and time since surgery (49.4 vs. 53.3 months). In the weighted 12-month cohort, mean BMI was 39.5 (SD = 8.0) and 39.1 (SD = 8.4) for users and non-users, respectively, with similar nadir BMI (31.5 vs. 31.4) and time since surgery (50.4 vs. 52.6 months). Weighted baseline characteristics are presented in **Table 1**, with detailed pre- and post-weighting comparisons provided in Supplementary **eTables 3 and 4.**

**Table 1.** Baseline characteristics of GLP-1RA initiators and non-initiators after matching and IPTW.

|  | At 6 months |  |  | At 12 months |  |  |
| --- | --- | --- | --- | --- | --- | --- |
| Characteristics | GLP-1RA Users, No(%) (n=68) | GLP-1RA Non-Users, No(%) (n=131) | SMD | GLP-1RA Users, No(%) (n=72) | GLP-1RA Non-Users, No(%) (n=139) | SMD |
| **Age (%)** |  |  |  |  |  |  |
| 18-40 | 12 (19.53%) | 26 (19.05%) | 0.0120 | 12 (18.09%) | 27 (19.04%) | -0.0243 |
| 40-65 | 50 (65.36%) | 75 (61.05%) | 0.0895 | 53 (67.01%) | 77 (58.79%) | 0.1707 |
| >=65 | 9 (15.11%) | 30 (19.90%) | -0.1262 | 9 (14.89%) | 35 (22.17%) | -0.1880 |
| **Sex (%)** |  |  |  |  |  |  |
| Female | 60 (83.75%) | 99 (79.02%) | 0.1217 | 62 (79.99%) | 104 (77.66%) | 0.0570 |
| Male | 11 (16.25%) | 32 (20.98%) | -0.1217 | 12 (20.01%) | 35 (22.34%) | -0.0570 |
| **Race Ethnicity (%)** |  |  |  |  |  |  |
| Asian | 0 (0.00%) | 2 (1.38%) | -0.1673 | 0 (0.00%) | 2 (1.41%) | -0.1689 |
| Black | 31 (40.19%) | 52 (41.28%) | -0.0220 | 32 (42.33%) | 55 (42.09%) | 0.0049 |
| Hispanic | 6 (7.55%) | 9 (8.88%) | -0.0484 | 6 (6.88%) | 8 (5.42%) | 0.0609 |
| Non-Hispanic Others | 0 (0.00%) | 0 (0.00%) | 0.0000 | 0 (0.00%) | 0 (0.00%) | 0.0000 |
| Unknown | 4 (4.63%) | 6 (4.22%) | 0.0201 | 4 (4.21%) | 7 (4.52%) | -0.0151 |
| White | 30 (47.62%) | 62 (44.24%) | 0.0678 | 32 (46.58%) | 67 (46.57%) | 0.0002 |
| **Insurance Coverage (%)** |  |  |  |  |  |  |
| MEDICAID | 1 (2.12%) | 8 (4.85%) | -0.1497 | 1 (1.74%) | 8 (4.77%) | -0.1717 |
| MEDICARE | 6 (8.49%) | 27 (16.66%) | -0.2482 | 7 (8.53%) | 32 (18.85%) | -0.3036 |
| NO_PAYMENT | 1 (1.01%) | 2 (1.18%) | -0.0161 | 1 (0.88%) | 2 (1.14%) | -0.0268 |
| OTHER | 0 (0.00%) | 2 (1.12%) | -0.1504 | 0 (0.000%) | 2 (1.08%) | -0.1477 |
| PRIVATE | 34 (51.93%) | 61 (48.08%) | 0.0770 | 34 (52.36%) | 64 (48.29%) | 0.0815 |
| UNKNOWN | 29 (36.46%) | 31 (28.11%) | 0.1791 | 31 (36.5%) | 31 (25.87%) | 0.2310 |
| **Smoking Status (%)** |  |  |  |  |  |  |
| Current Smoker | 0 (0.00%) | 2 (1.11%) | -0.1500 | 0 (0.00%) | 2 (1.08%) | -0.1475 |
| Former Smoker | 2 (3.00%) | 1 (3.27%) | -0.0157 | 2 (2.79%) | 0 (0.00%) | 0.2396 |
| Never Smoker | 7 (7.20%) | 13 (9.15%) | -0.0713 | 7 (6.49%) | 15 (9.98%) | -0.1272 |
| Unknown | 62 (89.80%) | 115 (86.47%) | 0.1034 | 65 (90.72%) | 122 (88.95%) | 0.0588 |
| **BMI Group (%)** |  |  |  |  |  |  |
| 25-30 | 3 (5.81%) | 12 (7.77%) | -0.0781 | 4 (11.22%) | 15 (9.56%) | 0.0545 |
| 30-35 | 21 (27.06%) | 29 (22.52%) | 0.1053 | 22 (25.55%) | 31 (23.55%) | 0.0465 |
| 35-40 | 9 (14.73%) | 30 (19.50%) | -0.1269 | 10 (15.45%) | 32 (20.50%) | -0.1319 |
| <25 | 0 (0.00%) | 6 (3.54%) | -0.2709 | 0 (0.00%) | 6 (3.41%) | -0.2656 |
| >=40 | 38 (52.40%) | 54 (46.67%) | 0.1149 | 38 (47.79%) | 55 (42.99%) | 0.0965 |
| **Weight information (N (std))** |  |  |  |  |  |  |
| Weight closest to index date | 247.42 (45.61) | 244.92 (54.62) | 0.0497 | 241.8 (47.41) | 241.29 (53.83) | 0.0101 |
| Weight mean (in baseline period) | 243.51 (45.39) | 242.05 (55.08) | 0.0290 | 238.45 (46.81) | 238.39 (54.17) | 0.0011 |
| BMI closest to index date | 40.33 (7.67) | 39.66 (8.49) | 0.0836 | 39.51 (7.99) | 39.05 (8.36) | 0.0560 |
| BMI mean (in baseline period) | 39.62 (7.63) | 39.23 (8.51) | 0.0485 | 38.89 (7.91) | 38.63 (8.39) | 0.0324 |
| Nadir BMI (in baseline period) | 32.13 (6.97) | 31.77 (7.33) | 0.0496 | 31.54 (7.04) | 31.42 (7.22) | 0.0165 |
| Nadir Weight (in baseline period) | 201.78 (39.89) | 197.48 (47.3) | 0.0982 | 196.94 (41.58) | 195.31 (46.36) | 0.0370 |
| Bariatric Surgery to Index Date (months) | 49.42 (20.19) | 53.33 (21.67) | -0.1867 | 50.41 (21.55) | 52.57 (21.37) | -0.1004 |
| Follow-up time (months) | 19.53 (16.21) | 14.9 (12.48) | 0.3651 | 21.4 (15.79) | 15.03 (12.37) | 0.3761 |
| **Comorbidities (%)** |  |  |  |  |  |  |
| COPD | 3 (7.39%) | 14 (9.14%) | -0.0637 | 3 (3.56%) | 17 (10.71%) | -0.2803 |
| Asthma | 8 (15.48%) | 25 (17.16%) | -0.0456 | 9 (15.34%) | 25 (17.27%) | -0.0522 |
| Hypertension | 48 (67.16%) | 87 (68.25%) | -0.0233 | 51 (69.53%) | 90 (66.72%) | 0.0603 |
| Dyslipidemia | 33 (45.53%) | 57 (43.61%) | 0.0386 | 34 (39.62%) | 57 (40.59%) | -0.0198 |
| Type 2 Diabetes (T2D) | 28 (35.78%) | 42 (35.93%) | -0.0030 | 30 (37.79%) | 42 (32.99%) | 0.1004 |
| Rheumatoid Arthritis/Osteoarthritis | 21 (28.19%) | 55 (36.63%) | -0.1812 | 22 (30.87%) | 57 (37.28%) | -0.1355 |
| Sleep apnea | 22 (29.47%) | 51 (34.70%) | -0.1121 | 23 (31.88%) | 53 (35.33%) | -0.0731 |
| Stroke | 1 (1.82%) | 3 (2.31%) | -0.0345 | 1 (1.67%) | 3 (2.23%) | -0.0404 |
| Myocardial infarction | 1 (1.82%) | 5 (3.34%) | -0.0959 | 1 (1.67%) | 5 (3.25%) | -0.1022 |
| Coronary Artery Disease | 9 (12.21%) | 19 (13.16%) | -0.0283 | 9 (11.16%) | 20 (13.42%) | -0.0687 |
| GERD | 28 (44.33%) | 64 (45.14%) | -0.0163 | 29 (46.85%) | 65 (45.07%) | 0.0357 |
| Depression | 16 (22.31%) | 34 (22.62%) | -0.0074 | 17 (21.91%) | 35 (23.28%) | -0.0327 |

After applying IPTW, all covariates were well balanced between groups, with absolute SMDs <0.1

### Recurrence Weight Lost

Among patients who experienced at least 10% recurrent weight after MBS, GLP-1 RA users lost significantly more recurrent weight than non-users at both time points. At 6 months, GLP-1 RA users achieved a mean recurrent weight loss of 9.6% compared to 4.3% in non-users (p = 0.005; 95%: CI 0.21–1.17). At 12 months, users achieved 14.8% recurrent weight loss compared to 8.1% in non-users (p = 0.017; 95%: CI 0.10–1.01). Both groups demonstrated progressive improvement from 6 to 12 months, with GLP-1 RA users maintaining a significant advantage.

### Subgroup Analyses by GLP-1 RA Type

Tirzepatide users achieved the greatest reductions in recurrent weight. At 6 months, tirzepatide users had a mean recurrent weight loss of 25.1% compared with a mean of 5.4% in non-users (p < 0.001; 95% CI: 0.95–1.85). At 12 months, tirzepatide users had a mean recurrent weight loss of 34.4% compared with 8.8% in non-users (p < 0.001; 95% CI: 0.83– 1.73).

Liraglutide users had a mean recurrent weight loss of 9.5% at 6 months compared with 5.1% in non-users, though this difference was not statistically significant (p = 0.132; 95% CI: −0.17–1.25). At 12 months, liraglutide users had a mean recurrent weight loss of 18.3% compared with 8.4% in non-users (p = 0.004; 95% CI: 0.24–1.20).

Semaglutide users had a mean recurrent weight loss of 6.5% at 6 months compared with 5.0% in non-users (p = 0.418; 95% CI: −0.32–0.77), and a mean loss of 8.7% at 12 months compared with 8.3% in non-users (p = 0.880; 95% CI: −0.51–0.60), with no statistically significant differences at either time point.

Dulaglutide users had a mean recurrent weight loss of 4.9% at 6 months compared with 5.2% in non-users (p = 0.897; 95% CI: –0.75–0.66). At 12 months, dulaglutide users had a mean recurrent weight loss of 6.5% compared with 8.8% in non-users (p = 0.469; 95% CI: −0.98–0.45), with no statistically significant differences observed. Subgroup analyses could not be performed for exenatide and lixisenatide due to insufficient sample sizes.

### Subgroup Analyses by Diabetes Status

Among patients with type 2 diabetes, GLP-1 RA users experienced greater recurrent weight loss at 6 months, achieving a mean loss of 13.6% compared with 4.6% in non-users (p = 0.007; 95% CI: 0.26–1.65). At 12 months, users had a mean recurrent weight loss of 15.2% compared with 10.7% in non-users, although this difference was not statistically significant (p = 0.37; 95% CI: –0.39–1.03).

Among patients without type 2 diabetes, recurrent weight loss at 6 months was a mean of 6.4% for users and 4.9% for non-users (p = 0.466; 95% CI: –0.38–0.83), indicating no statistically significant difference. By 12 months, users demonstrated significantly greater recurrent weight loss, achieving a mean of 14.6% compared with 7.1% in non-users (p = 0.016; 95% CI: 0.12–1.19).

### Safety Outcomes

GLP-1 RA users and non-users experienced similar rates of gastrointestinal adverse events at both 6 and 12 months. At 6 months, abdominal pain occurred in 16.2% of users compared with 26.7% of non-users, nausea with or without vomiting in 11.8% vs. 12.2%, and GERD in 32.4% vs. 36.6%, respectively. By 12 months, abdominal pain (15.3% vs. 25.2%) and nausea (11.1% vs. 11.5%) remained comparable between groups, while GERD persisted at similar levels (31.9% vs. 34.5%). The composite safety outcome occurred in 47.1% of users versus 56.5% of non-users at 6 months, and 45.8% vs. 53.2% at 12 months. Additional adverse events, including bowel obstruction, internal hernia, incisional hernia, gastrojejunal ulcer, gastrointestinal hemorrhage, gastric or duodenal fistula, and dehydration, occurred infrequently in both cohorts. The majority affected fewer than 10% of participants at each assessment period, with no discernible differences observed between GLP-1 RA users and non-users.

### Comparative Risk Analysis

After adjustment for baseline covariates, GLP-1 RA use was not associated with increased risk of adverse events at either time point. At 6 months, adjusted hazard ratios comparing users to non-users were (HR = 1.21, 95% CI: 0.76–1.91) for abdominal pain, (HR = 1.24, 95% CI: 0.69–2.23) for nausea with or without vomiting, and (HR = 1.18, 95% CI: 0.81–1.71) for GERD. At 12 months, corresponding HRs were (HR = 0.70, 95% CI: 0.48–1.02), (HR = 1.14, 95% CI: 0.70–1.84), and (HR = 0.79, 95% CI: 0.57–1.10), respectively. The composite safety outcome showed similar risk at 6 months (HR = 1.24, 95% CI: 0.92–1.68) with a trend toward lower risk at 12 months (HR = 0.80, 95% CI: 0.61–1.05). None of these associations reached statistical significance (p > 0.05 for all). Additional outcomes including gastroparesis, dehydration, bowel obstruction, internal hernia, incisional hernia, gastric or duodenal fistula, post-bariatric surgery symptoms, and general complications of bariatric surgery occurred infrequently and could not be reliably modeled using Cox regression due to small sample sizes.

## Discussion

In this population-level cohort study of patients who experienced weight recurrence following MBS, the initiation of GLP-1 RA, particularly tirzepatide and liraglutide, was associated with significant decreases in weight without evidence of increased risk of adverse events. These findings provide real-world evidence supporting the use of GLP-1 RA as safe and effective treatment options for the management of weight recurrence following MBS across diverse patient populations.

The magnitude of weight recurrence lost while with GLP-1 RA use observed in this study aligns with previous single-institution literature evaluating the efficacy of these medications following MBS.^16,18,21^ Previous reports have suggested that the use of GLP-1 RA prior to MBS may diminish post-operative weight loss, highlighting concerns that combining MBS and GLP-1 RA therapies may diminish the weight loss impact of one treatment.^23^ However, our results highlight a clear benefit for the use of a range of GLP-1 RA for the treatment of weight recurrence. Furthermore, the progressive improvement in weight loss from 6 to 12 months observed in this study suggests sustained therapeutic benefit with continued treatment, addressing potential concerns about diminishing efficacy over time. This sustained weight loss benefit is particularly relevant given that weight regain typically begins 12-24 months following MBS and can compromise the metabolic benefits achieved.^8^ Therefore, the adjuvant use of these GLP-1 RA represents an important consideration for patients with weight recurrence as the substantial weight loss observed may help restore improvements in obesity-related comorbidities which may deteriorate as patients gain weight.

Evaluation of patient and medication subgroups revealed important nuances regarding weight recurrence treatment efficacy. Tirzepatide users experienced the greatest reduction in recurrent weight at 6- and 12-months post treatment initiation, suggesting strong real-world effectiveness in the post-MBS population. Furthermore, while liraglutide use demonstrated significant weight recurrence recovery at 12, semaglutide and dulaglutide showed non-significant trends. These findings differ from recent comparative studies supporting the effectiveness of semaglutide for the treatment of weight recurrence.^24^ The differences may reflect variations in patient populations, dosing regimens, treatment adherence as has been reported previously with adjuvant semaglutide.^17^ Interestingly, our analysis by diabetes status revealed that patients with type 2 diabetes showed significant early benefit at 6 months (13.6% vs. 4.6%, p = 0.007), while those without diabetes demonstrated greater separation at 12 months (14.6% vs. 7.1%, p = 0.016). Taken together, these findings suggest that GLP-1 RA may benefit diverse patient populations, though optimal agent selection and timing may vary based on individual characteristics.

The safety profile results within this study revealed no evidence of worse outcomes associated with GLP-1 RA use following MBS. These results match previous reports demonstrating acceptable safety when GLP-1 RA are used in the perioperative period.^16^ While concerns regarding GI-related adverse events associated with GLP-1 RA use may limit the use of these therapies following MBS, recent multidisciplinary consensus recommendations emphasize that GI adverse events can be effectively managed through dose titration and patient education.^25^ Furthermore, our findings of comparable rates of complications associated with MSB, including bowel obstruction, internal hernia, and gastrointestinal hemorrhage, between groups further support the safe use of GLP-1 RA in this patient population.

This work has several limitations. First, the retrospective nature of our study design introduces potential for unmeasured confounding which may not have been completely mitigated with the use of propensity score matching and inverse probability weighting. Second, selection bias may exist, as patients prescribed GLP-1 RA may have differed from non-users in motivation, adherence to follow-up, or other unmeasured characteristics that influence treatment outcomes. Third, the relatively small sample size in certain subgroup analyses, particularly for semaglutide may limited statistical power to detect meaningful differences. Fourth, we could not assess treatment adherence, discontinuation rates, or durability of weight loss beyond 12 months, all of which are important clinical considerations. Finally, the definition of weight recurrence as ≥10% increase from nadir weight, while clinically meaningful, may not capture patients with lesser degrees of regain who might also benefit from GLP-1 RA therapy. Despite these limitations, this study provides valuable real-world evidence to guide clinical decision-making for this common post-bariatric challenge.

## Conclusion

This population-level cohort study demonstrates that GLP-1 RA use for the treatment of weight recurrence following MBS is both effective and safe. These findings support the integration of GLP-1 RA therapies into the treatment algorithm for weight recurrence following MBS across diverse patient populations. Further work should focus on identifying ideal timing and duration of GLP-1 RA therapy as well as evaluating the long-term durability of weight loss to further refine clinical decision making regarding these treatments.

## Data Availability

Access to OneFlorida+ data is available to eligible researchers through a formal application and approval process administered by the OneFlorida+ Research Consortium. Investigators may request access by submitting a proposal to the OneFlorida+ Data Trust Review Committee. Additional information is available at the OneFlorida+ website.
The analytic code and variable definitions used in this study are available from the corresponding author upon reasonable request and pending institutional approvals.

