## Supplemental material for "Real-World Effectiveness and Safety of GLP-1 Receptor Agonists in Patients with Weight Recurrence After Bariatric Surgery"

**Table 1.** Bariatric surgery identification codes.

| **Surgery Category** | **ICD-9** | **ICD-10** | **CPT/HCPCS** |
| --- | --- | --- | --- |
| Gastric Bypass (variants of gastric bypass surgery (Roux-en-Y), differing by approach (open vs endoscopic), device use, and specific bypass configuration) | 44.31, 43.7, 44.38, 44.39 | 0D16079, 0D1607A, 0D1607B, 0D1607L, 0D160J9, 0D160JA, 0D160JB, 0D160JL, 0D160K9, 0D160KA, 0D160KB, 0D160KL, 0D160Z9, 0D160ZA, 0D160ZB, 0D160ZL, 0D16479, 0D1647A, 0D1647B, 0D1647L, 0D164J9, 0D164JA, 0D164JB, 0D164JL, 0D164K9, 0D164KA, 0D164KB, 0D164KL, 0D164Z9, 0D164ZA, 0D164ZB, 0D164ZL, 0D16078, 0D1687A, 0D168JA, 0D168K9, 0D168KA, 0D168ZA, 0D168Z9, 0D16879, 0D168J9, 0DB60ZZ | 43633, 43644, 43645, 43846, 43847, 43844 |
| Sleeve Gastrectomy (open and laparoscopic) | 43.82, 43.89, 44.68, 44.69 | 0DB60Z3,  0DB63Z3,  0DB63ZZ, 0DB67Z3, 0DB67ZZ, 0DQ60ZZ, 0DB64Z3, 0DQ64ZZ, 0DQ67ZZ, 0DQ63ZZ | 43775, 43843, 43842 |
| Open Vertical-Banded Gastroplasty (VBG) | 44.68 | 0DQ64ZZ | 43842 |
| Biliopancreatic Diversion / Gastric Reduction Duodenal Switch (BPD/GRDS) | 45.91, 45.51, 43.89 | 0D190Z9, 0DB60ZZ, 0DB80ZZ | 43845 |
| Laparoscopic Adjustable Gastric Band (AGB) | 44.95 | 0DV64CZ | 43770, S2082 |
| Revisional Bariatric Surgery | 44.96, 44.5 | 0DW643Z, 0DW64CZ | 43281, 43282, 43332, 43333, 43334, 43335, 43336, 43337, 43771, 43772, 43773, 43774, 43848, 43850, 43855, 43860, 43865, 43886, 43887, 43888 |

**Table 2.** Safety outcome identification codes.

| **Complication** | **ICD-9** | **ICD-10** |
| --- | --- | --- |
| Bowel obstruction | 560.1,560.2,560.8,560.9 | K56.0,K56.2,K56.5,K56.7,K56.9 |
| Internal hernia | 552.9,551.9,553.9 | K46.0,K46.1,K46.9 |
| Incisional Hernia | 551.21,552.21,553.21 | K43.1,K43.0,K43.2 |
| Fistula of stomach/duodenum | 537.4 | K31.6 |
| Post-gastric surgery symptoms | 564.2 | K91.1 |
| Complications of bariatric surgery | 539.0,539.01,539.0,539.81,539.89 | K95.0,K95.0,K95.01,K95.09,K95.8,K95.81,K95.89 |
| Abdominal pain | 789.0,789.01,789.02,789.03,789.04,789.05,789.06,789.07,789.08,789.09,789.6,789.60,789.61,789.62,789.63,789.64,789.65,789.66,789.67,789.69 | R10,R10.0,R10.1,R10.3,R10.8R10.9 |
| Gastrojejunal Ulcer | 534.0,534.1,534.2,534.3,534.4,534.5,534.6,534.7,534.9 | K28.0,K28.1,K28.2,K28.3,K28.4,K28,5,K28.6,K28.7,K28.9 |
| Cholelithiasis/ cholecystitis | 574.0,574.1,574.2,575.0,575.10,575.11,575.12,575.2,575.3,575.4,575.5,575.9 | K81.0,K81.1,K81.2,K81.9,K82.0,K821.1,K82.2,K82.3,K82.A1,K82.A2,K82.9,K80.00,K80.01,K80.1,K80.10,K80.11,K80.12,K80.13,K80.18,K80.19,K80.20,K80.21,K80.80,K80.81 |
| Cholangitis/ Choledocholithiasis | 574.4,574.5,574.6,574.7,574.8,574.9,576.1,576.2,576.3,576.4 | K75.0,K80.30,K80.31,K80.32,K80.33,K80.34,K80.35,K80.36,K80.37,K80.4,K80.40,K80.41,K80.42,K80.43,K80.44,K80.45,K80.46,K80.47,K80.5,K80.50,K80.51,K83.0,K83.09,K83.1,K83.2,K83.3,K80.6,K80.60,K80.61,K80.62,K80.63,K80.64,K80.65,K80.66,K80.67,K80.70,K80.71 |
| GI hemorrhage | 578.0,578.1,578.9 | K92.0,K92.1,K92.2 |
| Dehydration | 276.50,276.52,276.0,276.1 | E86.0 |
| Nausea with/ without vomiting | 787.01,787.02,787.0 | R11.0,R11.2 |
| GERD | 530.81,530.11 | K21,K21.0,K21.00,K21.01,K21.9 |
| Gastroparesis | 536.3 | K31.84 |

**Table 3.** Baseline characteristics of GLP-1RA Users vs GLP-1RA Non-Users cohort before and after IPTW – 6 months follow-up weight regain lost with 10 % thresholds

|  | **Before IPTW** | | | **After IPTW** | | |
| --- | --- | --- | --- | --- | --- | --- |
| **Characteristics** | **GLP-1RA Users, No(%) (n=71)** | **GLP-1RA Non-Users, No(%) (n=134)** | **SMD** | **GLP-1RA Users, No(%) (n=68)** | **GLP-1RA Non-Users, No(%) (n=131)** | **SMD** |
| **Age (%)** | | | | | | |
| 18-40 | 12 (16.90%) | 26 (19.40%) | -0.0641 | 12 (19.53%) | 26 (19.05%) | 0.0120 |
| 40-65 | 50 (70.42%) | 77 (57.46%) | 0.2678 | 50 (65.36%) | 75 (61.05%) | 0.0895 |
| >=65 | 9 (12.68%) | 31 (23.13%) | -0.2647 | 9 (15.11%) | 30 (19.90%) | -0.1262 |
| **Sex (%)** | | | | | | |
| Female | 60 (84.51%) | 100 (74.63%) | 0.2391 | 60 (83.75%) | 99 (79.02%) | 0.1217 |
| Male | 11 (15.49%) | 34 (25.37%) | -0.2391 | 11 (16.25%) | 32 (20.98%) | -0.1217 |
| **Race Ethnicity (%)** | | | | | | |
| Asian | 0 (0.00%) | 2 (1.49%) | -0.1515 | 0 (0.00%) | 2 (1.38%) | -0.1673 |
| Black | 31 (43.66%) | 53 (39.55%) | 0.0832 | 31 (40.19%) | 52 (41.28%) | -0.0220 |
| Hispanic | 6 (8.45%) | 9 (6.72%) | 0.0663 | 6 (7.55%) | 9 (8.88%) | -0.0484 |
| Non Hispanic Others | 0 (0.00%) | 0 (0.00%) | 0.0000 | 0 (0.00%) | 0 (0.00%) | 0.0000 |
| Unknown | 4 (5.63%) | 6 (4.48%) | 0.0534 | 4 (4.63%) | 6 (4.22%) | 0.0201 |
| White | 30 (42.25%) | 64 (47.76%) | -0.1101 | 30 (47.62%) | 62 (44.24%) | 0.0678 |
| **Insurance Coverage (%)** | | | | | | |
| MEDICAID | 1 (1.41%) | 8 (5.97%) | -0.2228 | 1 (2.12%) | 8 (4.85%) | -0.1497 |
| MEDICARE | 6 (8.45%) | 30 (22.39%) | -0.3702 | 6 (8.49%) | 27 (16.66%) | -0.2482 |
| NO_PAYMENT | 1 (1.41%) | 2 (1.49%) | -0.0070 | 1 (1.01%) | 2 (1.18%) | -0.0161 |
| OTHER | 0 (0.00%) | 2 (1.49%) | -0.1515 | 0 (0.00%) | 2 (1.12%) | -0.1504 |
| PRIVATE | 34 (47.89%) | 61 (45.52%) | 0.0472 | 34 (51.93%) | 61 (48.08%) | 0.0770 |
| UNKNOWN | 29 (40.85%) | 31 (23.13%) | 0.3942 | 29 (36.46%) | 31 (28.11%) | 0.1791 |
| **Smoking Status (%)** | | | | | | |
| Current Smoker | 0 (0.00%) | 2 (1.49%) | -0.1515 | 0 (0.00%) | 2 (1.11%) | -0.1500 |
| Former Smoker | 2 (2.82%) | 1 (0.75%) | 0.1722 | 2 (3.00%) | 1 (3.27%) | -0.0157 |
| Never Smoker | 7 (9.86%) | 14 (10.45%) | -0.0193 | 7 (7.20%) | 13 (9.15%) | -0.0713 |
| Unknown | 62 (87.32%) | 117 (87.31%) | 0.0003 | 62 (89.80%) | 115 (86.47%) | 0.1034 |
| **BMI Group (%)** | | | | | | |
| 25-30 | 3 (4.23%) | 13 (9.70%) | -0.2041 | 3 (5.81%) | 12 (7.77%) | -0.0781 |
| 30-35 | 21 (29.58%) | 29 (21.64%) | 0.1846 | 21 (27.06%) | 29 (22.52%) | 0.1053 |
| 35-40 | 9 (12.68%) | 31 (23.13%) | -0.2647 | 9 (14.73%) | 30 (19.50%) | -0.1269 |
| <25 | 0 (0.00%) | 7 (5.22%) | -0.2890 | 0 (0.00%) | 6 (3.54%) | -0.2709 |
| >=40 | 38 (53.52%) | 54 (40.30%) | 0.2667 | 38 (52.40%) | 54 (46.67%) | 0.1149 |
| **Weight information (N (std))** | | | | | | |
| Weight closest to index date | 248.3 (47.72) | 237.34 (48.27) | 0.2269 | 247.42 (45.61) | 244.92 (54.62) | 0.0497 |
| Weight mean (in baseline period) | 244.51 (47.30) | 234.32 (48.77) | 0.2101 | 243.51 (45.39) | 242.05 (55.08) | 0.0290 |
| BMI closest to index date | 40.66 (7.86) | 38.34 (7.68) | 0.2976 | 40.33 (7.67) | 39.66 (8.49) | 0.0836 |
| BMI mean (in baseline period) | 39.94 (7.77) | 37.88 (7.73) | 0.2646 | 39.62 (7.63) | 39.23 (8.51) | 0.0485 |
| Nadir BMI (in baseline period) | 32.63 (7.24) | 30.71 (6.75) | 0.2761 | 32.13 (6.97) | 31.77 (7.33) | 0.0496 |
| Nadir Weight (in baseline period) | 203.62 (42.99) | 190.99 (41.33) | 0.2999 | 201.78 (39.89) | 197.48 (47.3) | 0.0982 |
| Bariatric Surgery to Index Date (months) | 49.24 (20.94) | 53.58 (22.11) | -0.1989 | 49.42 (20.19) | 53.33 (21.67) | -0.1867 |
| Follow-up time (months) | 19.4 (15.26) | 14.78 (12.4) | 0.3048 | 19.53 (16.21) | 14.9 (12.48) | 0.3651 |
| **Comorbidities (%)** | | | | | | |
| COPD | 3 (4.23%) | 17 (12.69%) | -0.2864 | 3 (7.39%) | 14 (9.14%) | -0.0637 |
| Asthma | 8 (11.27%) | 26 (19.40%) | -0.2188 | 8 (15.48%) | 25 (17.16%) | -0.0456 |
| Hypertension | 48 (67.61%) | 89 (66.42%) | 0.0251 | 48 (67.16%) | 87 (68.25%) | -0.0233 |
| Dyslipidemia | 33 (46.48%) | 57 (42.54%) | 0.0791 | 33 (45.53%) | 57 (43.61%) | 0.0386 |
| Type 2 Diabetes (T2D) | 28 (39.44%) | 44 (32.84%) | 0.1379 | 28 (35.78%) | 42 (35.93%) | -0.0030 |
| Rheumatoid Arthritis/Osteoarthritis (RA/OA) | 21 (29.58%) | 58 (43.28%) | -0.2828 | 21 (28.19%) | 55 (36.63%) | -0.1812 |
| Sleep apnea | 22 (30.99%) | 53 (39.55%) | -0.1776 | 22 (29.47%) | 51 (34.70%) | -0.1121 |
| Stroke | 1 (1.41%) | 3 (2.24%) | -0.0598 | 1 (1.82%) | 3 (2.31%) | -0.0345 |
| Myocardial infarction | 1 (1.41%) | 5 (3.73%) | -0.1374 | 1 (1.82%) | 5 (3.34%) | -0.0959 |
| Coronary Artery Disease | 9 (12.68%) | 19 (14.18%) | -0.0436 | 9 (12.21%) | 19 (13.16%) | -0.0283 |
| GERD | 28 (39.44%) | 65 (48.51%) | -0.1820 | 28 (44.33%) | 64 (45.14%) | -0.0163 |
| Depression | 16 (22.54%) | 35 (26.12%) | -0.0826 | 16 (22.31%) | 34 (22.62%) | -0.0074 |

**Table 4.** Baseline characteristics of GLP-1RA Users vs GLP-1RA Non-Users cohort before and after IPTW – 12 months follow-up weight regain lost with 10 % thresholds

|  | **Before IPTW** | | | **After IPTW** | | |
| --- | --- | --- | --- | --- | --- | --- |
| **Characteristics** | **GLP-1RA Users, No(%) (n=74)** | **GLP-1RA Non-Users, No(%) (n=143)** | **SMD** | **GLP-1RA Users, No(%) (n=72)** | **GLP-1RA Non-Users, No(%) (n=139)** | **SMD** |
| **Age (%)** | | | | | | |
| 18-40 | 12 (16.22%) | 27 (18.88%) | -0.0691 | 12 (18.09%) | 27 (19.04%) | -0.0243 |
| 40-65 | 53 (71.62%) | 80 (55.94%) | 0.3242 | 53 (67.01%) | 77 (58.79%) | 0.1707 |
| >=65 | 9 (12.16%) | 36 (25.17%) | -0.3232 | 9 (14.89%) | 35 (22.17%) | -0.1880 |
| **Sex (%)** | | | | | | |
| Female | 62 (83.78%) | 106 (74.13%) | 0.2313 | 62 (79.99%) | 104 (77.66%) | 0.0570 |
| Male | 12 (16.22%) | 37 (25.87%) | -0.2313 | 12 (20.01%) | 35 (22.34%) | -0.0570 |
| **Race Ethnicity (%)** | | | | | | |
| Asian | 0 (0.00%) | 2 (1.40%) | -0.1460 | 0 (0.00%) | 2 (1.41%) | -0.1689 |
| Black | 32 (43.24%) | 56 (39.16%) | 0.0828 | 32 (42.33%) | 55 (42.09%) | 0.0049 |
| Hispanic | 6 (8.11%) | 9 (6.29%) | 0.0712 | 6 (6.88%) | 8 (5.42%) | 0.0609 |
| Non Hispanic Others | 0 (0.00%) | 0 (0.00%) | 0.0000 | 0 (0.00%) | 0 (0.00%) | 0.0000 |
| Unknown | 4 (5.41%) | 7 (4.90%) | 0.0232 | 4 (4.21%) | 7 (4.52%) | -0.0151 |
| White | 32 (43.24%) | 69 (48.25%) | -0.1001 | 32 (46.58%) | 67 (46.57%) | 0.0002 |
| **Insurance Coverage (%)** | | | | | | |
| MEDICAID | 1 (1.35%) | 8 (5.59%) | -0.2129 | 1 (1.74%) | 8 (4.77%) | -0.1717 |
| MEDICARE | 7 (9.46%) | 35 (24.48%) | -0.3846 | 7 (8.53%) | 32 (18.85%) | -0.3036 |
| NO_PAYMENT | 1 (1.35%) | 2 (1.40%) | -0.0040 | 1 (0.88%) | 2 (1.14%) | -0.0268 |
| OTHER | 0 (0.00%) | 2 (1.40%) | -0.1460 | 0 (0.000%) | 2 (1.08%) | -0.1477 |
| PRIVATE | 34 (45.95%) | 64 (44.76%) | 0.0238 | 34 (52.36%) | 64 (48.29%) | 0.0815 |
| UNKNOWN | 31 (41.89%) | 32 (22.38%) | 0.4371 | 31 (36.5%) | 31 (25.87%) | 0.2310 |
| **Smoking Status (%)** | | | | | | |
| Current Smoker | 0 (0.00%) | 2 (1.40%) | -0.1460 | 0 (0.00%) | 2 (1.08%) | -0.1475 |
| Former Smoker | 2 (2.70%) | 1 (0.70%) | 0.1714 | 2 (2.79%) | 0 (0.00%) | 0.2396 |
| Never Smoker | 7 (9.46%) | 15 (10.49%) | -0.0340 | 7 (6.49%) | 15 (9.98%) | -0.1272 |
| Unknown | 65 (87.84%) | 125 (87.41%) | 0.0128 | 65 (90.72%) | 122 (88.95%) | 0.0588 |
| **BMI Group (%)** | | | | | | |
| 25-30 | 4 (5.41%) | 15 (10.49%) | -0.1797 | 4 (11.22%) | 15 (9.56%) | 0.0545 |
| 30-35 | 22 (29.73%) | 31 (21.68%) | 0.1873 | 22 (25.55%) | 31 (23.55%) | 0.0465 |
| 35-40 | 10 (13.51%) | 34 (23.78%) | -0.2560 | 10 (15.45%) | 32 (20.50%) | -0.1319 |
| <25 | 0 (0.00%) | 7 (4.90%) | -0.2782 | 0 (0.00%) | 6 (3.41%) | -0.2656 |
| >=40 | 38 (51.35%) | 56 (39.16%) | 0.2466 | 38 (47.79%) | 55 (42.99%) | 0.0965 |
| **Weight information (N (std))** | | | | | | |
| Weight closest to index date | 246.58 (47.5) | 236.9 (48.52) | 0.2001 | 241.8 (47.41) | 241.29 (53.83) | 0.0101 |
| Weight mean (in baseline period) | 242.8 (47.09) | 233.93 (48.72) | 0.1834 | 238.45 (46.81) | 238.39 (54.17) | 0.0011 |
| BMI closest to index date | 40.38 (7.89) | 38.18 (7.6) | 0.2842 | 39.51 (7.99) | 39.05 (8.36) | 0.0560 |
| BMI mean (in baseline period) | 39.67 (7.79) | 37.74 (7.64) | 0.2497 | 38.89 (7.91) | 38.63 (8.39) | 0.0324 |
| Nadir BMI (in baseline period) | 32.4 (7.19) | 30.73 (6.59) | 0.2452 | 31.54 (7.04) | 31.42 (7.22) | 0.0165 |
| Nadir Weight (in baseline period) | 201.81 (43.12) | 191.42 (40.63) | 0.2492 | 196.94 (41.58) | 195.31 (46.36) | 0.0370 |
| Bariatric Surgery to Index Date (months) | 50.26 (21.45) | 53.35 (22.11) | -0.1406 | 50.41 (21.55) | 52.57 (21.37) | -0.1004 |
| Follow-up time (months) | 19.47 (15.25) | 14.93 (12.25) | 0.3111 | 21.4 (15.79) | 15.03 (12.37) | 0.3761 |
| **Comorbidities (%)** | | | | | | |
| COPD | 3 (4.05%) | 20 (13.99%) | -0.3250 | 3 (3.56%) | 17 (10.71%) | -0.2803 |
| Asthma | 9 (12.16%) | 26 (18.18%) | -0.1634 | 9 (15.34%) | 25 (17.27%) | -0.0522 |
| Hypertension | 51 (68.92%) | 93 (65.03%) | 0.0819 | 51 (69.53%) | 90 (66.72%) | 0.0603 |
| Dyslipidemia | 34 (45.95%) | 59 (41.26%) | 0.0944 | 34 (39.62%) | 57 (40.59%) | -0.0198 |
| Type 2 Diabetes (T2D) | 30 (40.54%) | 44 (30.77%) | 0.2062 | 30 (37.79%) | 42 (32.99%) | 0.1004 |
| Rheumatoid Arthritis/Osteoarthritis (RA/OA) | 22 (29.73%) | 60 (41.96%) | -0.2529 | 22 (30.87%) | 57 (37.28%) | -0.1355 |
| Sleep apnea | 23 (31.08%) | 55 (38.46%) | -0.1535 | 23 (31.88%) | 53 (35.33%) | -0.0731 |
| Stroke | 1 (1.35%) | 3 (2.10%) | -0.0553 | 1 (1.67%) | 3 (2.23%) | -0.0404 |
| Myocardial infarction | 1 (1.35%) | 5 (3.50%) | -0.1305 | 1 (1.67%) | 5 (3.25%) | -0.1022 |
| Coronary Artery Disease | 9 (12.16%) | 21 (14.69%) | -0.0728 | 9 (11.16%) | 20 (13.42%) | -0.0687 |
| GERD | 29 (39.19%) | 67 (46.85%) | -0.1540 | 29 (46.85%) | 65 (45.07%) | 0.0357 |
| Depression | 17 (22.97%) | 36 (25.17%) | -0.0510 | 17 (21.91%) | 35 (23.28%) | -0.0327 |
